# Epidemiological Burden and Projections of Pulmonary Arterial Hypertension in China: An Analysis from the Global Burden of Disease Study 2021

**DOI:** 10.1101/2025.01.13.25320465

**Authors:** Shuoshuo Wei, Yonghui Han, Min Liu, Hanli Wang, Zhiwei Lu, Yusheng Cheng, Jun Guo, Lei Zha

## Abstract

**Objective:** Pulmonary arterial hypertension (PAH) is a severe condition with high morbidity and mortality, yet its epidemiology in China remains poorly understood. This study aims to assess the burden of PAH in China from 1990 to 2021 and project trends to 2050, providing insights for effective management and prevention strategies.

**Method:** Data from the Global Burden of Disease (GBD) Study 2021 were analyzed, focusing exclusively on PAH. Key metrics included incidence, prevalence, mortality, and disability-adjusted life years (DALYs), along with their corresponding age-standardized rates (ASRs). Average annual percent changes (AAPCs) were calculated using joinpoint regression analysis, and a Bayesian age-period-cohort model was employed to project trends through 2050.

**Results:** In 2021, there were 41,135 estimated prevalent cases of PAH (95% UI: 32,838.91 to 51,357.25) in China, with an age-standardized prevalence rate (ASPR) of 2.24 per 100,000. Females accounted for approximately 58% of cases. PAH caused 7,318 deaths (95% UI: 4,835.72 to 9,075.75; 3,683 males, 3,635 females), with an age-standardized death rate (ASDR) of 0.42 per 100,000. From 1990 to 2021, the ASPR increased slightly from 2.07 (95% UI: 1.68 to 2.54) to 2.24 (95% UI: 1.81 to 2.75), while the ASDR decreased from 0.61 (95% UI: 0.46 to 0.83) to 0.42 (95% UI: 0.28 to 0.51). Projections suggest declining trends in both ASPR and ASDR, although the overall prevalence of PAH is expected to rise over the next 30 years.

**Conclusion:** PAH imposes a significant disease burden in China, particularly among women and older adults. These findings highlight the urgent need to enhance diagnostic capabilities and develop improved treatment strategies to address this challenging condition in the Chinese population.

## Introduction

Pulmonary arterial hypertension (PAH), a severe and life-threatening subtype of pulmonary hypertension (PH), is characterized by significant remodeling of the pulmonary vasculature^1^. This remodeling process leads to a progressive increase in pulmonary vascular resistance, imposing a substantial workload on the right ventricle^1,2^. The resulting ventricular strain induces hypertrophy and remodeling, ultimately compromising cardiac function and contributing to disease-related morbidity and mortality^3^. Globally, PAH represents a significant public health challenge affecting all age groups^4^, substantially impacting patients’ quality of life and placing considerable burdens on healthcare systems and economies^5^.

PAH is the Group 1 subtype of pulmonary hypertension which encompasses various etiologies, including idiopathic origins, heritable connective tissue diseases, congenital heart defects, portal hypertension, HIV infection, drug and toxin exposure, and multifactorial causes. Idiopathic PAH (IPAH) represents the predominant subtype, accounting for 50-60% of cases^6^. The disease presents with nonspecific symptoms that develop insidiously, including exertional dyspnea, fatigue, chest pain, syncope, palpitations, and peripheral edema^7–9^. These manifestations significantly impact patients’ physical mobility and emotional well-being, compromising their health-related quality of life^10,11^. Despite advances in targeted therapies improving outcomes and quality of life, the five-year survival rate remains relatively low at approximately 60%^6,12^. Significant challenges persist, including ongoing disability and and extensive healthcare needs, which impose substantial financial burdens on patients, healthcare systems, and society^13,14,15^.

Global epidemiological data on PAH primarily derives from registry-based studies^6,14–17^. Prevalence rates vary geographically, ranging from 15 to 50 cases per million in the US and Europe^18^, with the US reporting approximately 25 cases per million^19^ and Central Australia showing a higher prevalence of 48 per million^20^. The Global Burden of Diseases (GBD) study recently provided comprehensive global data, reporting an age-standardized prevalence of 2.28 per 100,000 and an age-standardized mortality rate of 0.27 deaths per 100,000 population in 2021, with women and elderly populations disproportionately affected^21^. In China, the epidemiological landscape differs notably, with congenital heart disease (CHD)-associated PAH being most prevalent^22^, and a higher proportion of systemic lupus Erythematosus(SLE)-associated PAH in connective tissue diseases(CTD)^23^. However, comprehensive epidemiological data for China remains limited^24^. Available studies suggests lower incidence and survival rates compared to Western countries; however, the prevalence may be underestimated due to diagnostic challenges and underreported cases^23,25–27^.

With the comprehensive data provided by GBD 2021, which integrates multiple sources through advanced statistical modeling^28^, we have a unique opportunity to conduct an in-depth assessment of the PAH disease burden in China. Our analysis encompasses trends in prevalence, incidence, mortality, and disability-adjusted life years (DALYs) from 1990 to 2021, with projections extending to 2050, focusing exclusively on Group 1 PAH. This assessment aims to provide crucial insights for developing targeted prevention and treatment strategies that address the evolving healthcare needs of the Chinese population.

## Methods

### Overview

This study utilized data from the Global Burden of Disease (GBD) 2021 study, accessible at http://ghdx.healthdata.org/gbd-results-tool. GBD 2021, the project’s latest iteration, assessed the burden of 371 diseases, injuries and 88 risk factors for 204 countries and territories from 1990–2021^28^. The GBD database employs advanced statistical methods to address missing data and adjust for confounding factors. Detailed descriptions of the GBD study design and methodologies are available in the existing literature^28^. Furthermore, the GBD Study adheres to the Guidelines for Accurate and Transparent Health Estimates Reporting (GATHER) Statement ^29^.

### Data acquisition and download

The GBD study provides a comprehensive assessment of disease burden by cause, age, sex, year, and location, enabling direct comparisons across populations, time periods, and regions. This study specifically focused on the burden of PAH. From the GBD 2021 database, we extracted data on PAH for all ages, including age-standardized, age-specific, and sex-specific prevalence, incidence, mortality, and DALYs. DALYs were calculated as the sum of years of life lost (YLLs) and years lived with disability (YLDs), with 95% uncertainty intervals (UIs) derived from the 25th and 975th values of 1,000 statistical draws^30^.

The current hemodynamic definition of PAH includes three key criteria obtained through right heart catheterization: a resting mean pulmonary arterial pressure exceeding 20 mm Hg, a pulmonary capillary wedge pressure below 15 mm Hg, and a pulmonary vascular resistance of at least 2 Wood units^31^. In resource-limited settings, echocardiography is often used to estimate pulmonary arterial pressure, although this method can impact the sensitivity and specificity of the diagnosis. The GBD study applied a standardized case definition requiring a physician diagnosis of PAH, supported by evidence from either right heart catheterization or echocardiography. Vital registration records were mapped to the GBD cause hierarchy using specific ICD codes: 416.0 (ICD-9) and I27.0 (ICD-10). To ensure the most accurate and conservative estimates of PAH burden, the analysis was restricted to deaths coded as I27.0 after the introduction of I27.2, with all input data from affected locations prior to I27.2 implementation excluded. This study focused exclusively on Group 1 PAH, excluding other forms of pulmonary hypertension.

### Statistical analysis

A comprehensive assessment was conducted to quantify the burden of PAH in China in 2021, including prevalence, incidence, mortality, and DALYs. The analysis also examined the distribution of the disease burden across different age groups and genders. To evaluate time trends in prevalence, incidence, mortality, and DALY rates from 1990 to 2021, we calculated the average annual percent changes (AAPCs) with corresponding 95% confidence intervals (CIs) using joinpoint regression analysis^32^. We also employed age-period-cohort (APC) modeling to assess the associations between age, period, birth cohort, and PAH mortality^33^. APC and AAPC were considered statistically significant if the 95% CI did not overlap with zero and the *P* <0.05.

Bayesian Age-Period-Cohort (BAPC) models were used to project future trends, utilizing integrated nested Laplace approximations (INLA) for full Bayesian inference. Critical features of BAPC models include the generation of age-specific and age-standardized projected rates and the automatic incorporation of Poisson noise for predictive distributions ^34,35^. The preceding BAPC analysis is based on the R package "BAPC" (version 0.0.36) and "INLA" (version 24.02.09)^36^. All statistical analyses and visualizations were conducted using the R statistical software program (version 4.4.3) and JD_GBDR (V2.31, Jing ding Medical Technology Co., Ltd). *P*<0.05 was considered statistically significant.

## Results

### Burden of Pulmonary Arterial Hypertension in 2021

The burden of PAH was measured through incidence, prevalence, mortality, and DALYs, along with their age-standardized rates for all ages in 2021. In China, there were 9257 incidence cases (95%UI:7349.72,11507.74) of PAH, with an age-standard incidence rate (ASIR) of 0.50 per 100,000, comprising 4,569 males and 4,688 females (**Table 1**). The prevalence reached 41,135 cases (95% UI: 32,838.91,51,357.25) with an age-standardized prevalence rate (ASPR) of 2.24 per 100,000, with females accounting for nearly 58% of cases (**Table 2**). PAH contributes to 7318 deaths (95%UI:4835.72,9075.75) with an age-standard death rate (ASDR) of 0.42 per 100,000, including 3683 males and 3635 females (**Table 3**). The DALYs attributed to 150941 (95%UI: 99583.31,186503.40) with an age-standardized DALY rate of 8.95(95%UI:6.04,11.13) per 100,000 person-years (**Table 4**).

**Table 1.** presents the number of cases and age-standardized incidence rates (ASIR) of pulmonary arterial hypertension in China for the years 1990 and 2021, along with their temporal trends from 1990 to 2021.

|  | cases number |  | Incidence rate |  | EAPC,95%CI |
| --- | --- | --- | --- | --- | --- |
|  | 1990,95%UI | 2021,95%UI | 1990,95%UI | 2021,95%UI | ( 1990-2021 ) |
| Both* | 5125.86(4139.32,6211.42) | 9257.24(7349.72,11507.74) | 0.51(0.41,0.62) | 0.50(0.40,0.60) | -0.12(-0.17,-0.08) |
| male* | 2537.10(2041.82,3071.67) | 4568.97(3611.35,5721.01) | 0.50(0.40,0.60) | 0.49(0.40,0.60) | -0.03(-0.09,0.02) |
| female* | 2588.76(2094.71,3127.96) | 4688.27(3732.05,5864.27) | 0.52(0.42,0.63) | 0.50(0.40,0.61) | -0.21(-0.26,-0.17) |
| **Age\*\*** |  |  |  |  |  |
| <5 years | 176.44(131.22,237.53) | 121.76(90.00,163.41) | 0.16(0.12,0.21) | 0.16(0.12,0.21) | -0.04(-0.07,-0.01) |
| 5-9 years | 162.51(118.03,215.26) | 147.78(107.22,195.10) | 0.16(0.11,0.21) | 0.15(0.11,0.20) | -0.05(-0.09,-0.01) |
| 10-14 years | 158.27(93.34,247.58) | 130.84(76.82,203.65) | 0.16(0.09,0.24) | 0.15(0.09,0.24) | -0.07(-0.11,-0.02) |
| 15-19 years | 225.55(143.95,346.81) | 129.85(82.26,199.89) | 0.18(0.11,0.27) | 0.17(0.11,0.27) | -0.10(-0.16,-0.04) |
| 20-24 years | 296.07(189.33,437.69) | 160.68(101.82,238.42) | 0.22(0.14,0.33) | 0.22(0.14,0.33) | -0.12(-0.17,-0.07) |
| 25-29 years | 296.52(163.33,468.34) | 229.92(126.26,367.63) | 0.27(0.15,0.43) | 0.27(0.15,0.43) | -0.11(-0.15,-0.07) |
| 30-34 years | 300.47(193.36,442.31) | 408.11(260.97,602.43) | 0.34(0.22,0.50) | 0.34(0.22,0.50) | -0.09(-0.13,-0.05) |
| 35-39 years | 401.42(267.91, 583.04) | 457.30(303.38,668.28) | 0.44(0.29,0.64) | 0.43(0.29,0.63) | -0.09(-0.13,-0.05) |
| 40-44 years | 360.93(198.05,57.48) | 482.46(266.99,763.66) | 0.54(0.30,0.85) | 0.53(0.29,0.83) | -0.09(-0.13,-0.04) |
| 45-49 years | 350.49(219.298,51.27) | 732.86(457.18,1072.52) | 0.68(0.43,0.99) | 0.66(0.41,0.97) | -0.11(-0.16,-0.05) |
| 50-54 years | 408.95(272.40,598.81) | 1013.69(675.03,1485.14) | 0.86(0.57,1.26) | 0.84(0.56,1.23) | -0.13(-0.19,-0.07) |
| 55-59 years | 448.25(265.48,708.71) | 1113.16(659.85,1770.29) | 1.03(0.61,1.63) | 1.01(0.60,1.61) | -0.14(-0.19,-0.08) |
| 60-64 years | 443.14(291.02,648.88) | 892.68(590.18,1305.06) | 1.25(0.82,1.84) | 1.22(0.81,1.79) | -0.14(-0.18,-0.09) |
| 65-69 years | 414.14(296.59,559.20) | 1134.91(817.91,1527.88) | 1.52(1.09,2.05) | 1.48(1.07,1.99) | -0.14(-0.18,-0.09) |
| 70-74 years | 335.19(220.43,489.32) | 925.00(607.86,1346.72) | 1.78(1.17,2.60) | 1.74(1.14,2.53) | -0.14(-0.18,-0.09) |
| 75-79 years | 214.48(141.65,307.18) | 605.56(397.95,866.10) | 1.89(1.25,2.70) | 1.83(1.20,2.62) | -0.17(-0.21,-0.12) |
| 80-84 years | 96.67(69.00,132.30) | 347.82(248.73,475.47) | 1.83(1.30,2.50) | 1.76(1.26,2.40) | -0.22(-0.27,-0.16) |
| 85-89 years | 29.99(18.49,46.39) | 160.39(98.87,250.50) | 1.78(1.10,2.75) | 1.68(1.04,2.63) | -0.27(-0.32,-0.22) |
| 90-94 years | 5.61(3.36,8.82) | 50.54(29.73,79.20) | 1.83(1.10,2.87) | 1.72(1.01,2.70) | -0.29(-0.34,-0.24) |
| 95+ years | 0.80(0.41,1.46) | 11.94(6.03,21.77) | 1.98(1.00,3.60) | 0.87(0.94,3.41) | -0.27(-0.31,-0.23) |

Note:95% UI,95% uncertainty intervals. \*, age-standardized of incidence rate; \*\*, crude incidence rate in each age group, EAPC, estimated annual percentage change.

**Table 2.** presents the number of cases and age-standardized prevalence rates (ASPR) of pulmonary arterial hypertension in China for the years 1990 and 2021, along with their temporal trends from 1990 to 2021.

|  | cases number |  | Prevalence rate |  | EAPC,95%CI |
| --- | --- | --- | --- | --- | --- |
|  | 1990,95%UI | 2021,95%UI | 1990,95%UI | 2021,95%UI | ( 1990-2021 ) |
| Both* | 22027.91(17840.18,27321.46) | 41135.19(32838.91,51357.25) | 2.07(1.68,2.54) | 2.24(1.81,2.75) | 0.39(0.34,0.44) |
| male* | 9236.83(7510.66,11511.55) | 17482.09(13972.87,21744.92) | 1.70(1.37,2.09) | 1.91(1.54,2.34) | 0.56(0.51,0.62) |
| female* | 12791.08(10432.22,15798.13) | 23653.10(18864.64,29535.98) | 2.45(1.99,3.00) | 2.57(2.08,3.18) | 0.26(0.21,0.32) |
| Age* * |  |  |  |  |  |
| <5 years | 266.46(192.056,365.66) | 208.47(149.40,285.26) | 0.24(0.17,0.33) | 0.27(0.19,0.37) | 0.29(0.23,0.36) |
| 5-9 years | 500.44(360.48,656.99) | 493.40(355.25,650.62) | 0.48(0.35,0.63) | 0.52(0.37,0.68) | 0.34(0.31,0.38) |
| 10-14 years | 669.69(446.07,957.53) | 606.15(408.65,853.84) | 0.66(0.44,0.94) | 0.70(0.47,0.99) | 0.33(0.28,0.37) |
| 15-19 years | 1141.72(658.80,1756.51) | 712.923(418.45,1075.24) | 0.90(0.52,1.39) | 0.96(0.56,1.44) | 0.30(0.25,0.34) |
| 20-24 years | 1610.51(1067.98,2398.08) | 945.97(631.92,1386.88) | 1.22(0.81,1.82) | 1.29(0.86,1.90) | 0.31(0.26,0.36) |
| 25-29 years | 1743.19(1157.94,2567.82) | 1464.46(977.73,2140.53) | 1.59(1.05,2.34) | 1.69(1.13,2.48) | 0.33(0.29,0.38) |
| 30-34 years | 1761.80(1110.00,2628.85) | 2603.04(1670.21,3905.22) | 2.00(1.26,2.98) | 2.15(1.38,3.22) | 0.34(0.30,0.37) |
| 35-39 years | 2184.36(1496.04,3080.35) | 2734.65(1875.72,3865.12) | 2.39(1.64,3.37) | 2.58(1.77,3.65) | 0.36(0.32,0.39) |
| 40-44 years | 1818.06(1213.67,2575.10) | 2685.29(1806.42,3746.32) | 2.71(1.81,3.84) | 2.93(1.97,4.09) | 0.39(0.34,0.44) |
| 45-49 years | 1561.69(977.01,2331.54) | 3624.79(2305.10,5319.99) | 3.03(1.89,4.52) | 3.29(2.09,4.82) | 0.42(0.36,0.48) |
| 50-54 years | 1684.38(1150.43,2394.31) | 4642.16(3187.82,6592.07) | 3.53(2.41,5.02) | 3.84(2.64,5.45) | 0.44(0.38,0.50) |
| 55-59 years | 1790.39(1195.29,2550.84) | 4941.89(3345.14,7034.48) | 4.13(2.76,5.88) | 4.50(3.04,6.40) | 0.45(0.39,0.50) |
| 60-64 years | 1676.82(1061.18,2488.21) | 3777.42(2418.20,5571.49) | 4.75(3.00,7.04) | 5.17(3.31,7.63) | 0.44(0.39,0.48) |
| 65-69 years | 1466.71(1031.55,2033.45) | 4480.52(3147.24,6207.00) | 5.38(3.78,7.45) | 5.84(4.10,8.09) | 0.42(0.37,0.46) |
| 70-74 years | 1124.36(777.65,1602.87) | 3445.87(2394.75,4866.21) | 5.98(4.13,8.52) | 6.47(4.49,9.13) | 0.39(0.34,0.44) |
| 75-79 years | 691.21(432.86,1078.55) | 2180.29(1365.44,3332.07) | 6.07(3.80,9.48) | 6.58(4.12,10.06) | 0.42(0.35,0.49) |
| 80-84 years | 260.43(172.03,374.47) | 1063.65(704.08,1517.02) | 4.92(3.25,7.07) | 5.37(3.56,7.67) | 0.52(0.42,0.61) |
| 85-89 years | 65.00(39.45,101.55) | 403.86(246.95,625.85) | 3.85(2.34,6.02) | 4.24(2.59,6.57) | 0.54(0.45,0.64) |
| 90-94 years | 9.60(5.52,15.51) | 100.98(58.50,157.97) | 3.13(1.80,5.06) | 3.44(2.00,5.4) | 0.53(0.43,0.63) |
| 95+ years | 1.11(0.56,2.06) | 19.41(9.78,34.38) | 2.75(1.39,5.10) | 3.04(1.53,5.38) | 0.54(0.42,0.65) |

Note:95% UI,95% uncertainty intervals. \*, age-standardized prevalence rate; \*\*, crude incidence rate in each age group, EAPC, estimated annual percentage change.

Analysis of age-specific patterns revealed distinct distributions across different measures and metrics (**Figure 1**). The highest number of incident cases was observed in the 65-69 age group (1135 cases), while the peak crude incidence rate occurred in the 75-79 age group (1.83 per 100,000) (**Figure 1A**). For prevalence, the highest number of cases was recorded in the 55-59 age group (4,942 cases), while the highest crude prevalence rate was observed in the 75-79 age group (6.58 per 100,000) **(Figure 1B).** By contrast to prevalence, the greatest number of deaths occurred in the 80-84 age group (1,291 cases), while the highest crude death rate was observed in individuals aged 95+ years (23.13 per 100,000) (**Figure 1C**). The 70-74 age group had the largest number of DALYs (19236 cases), with the highest DALYs rate occurring in the 95+ age group (189.06 per 100,000) (**Figure 1D**).

**Figure 1.**
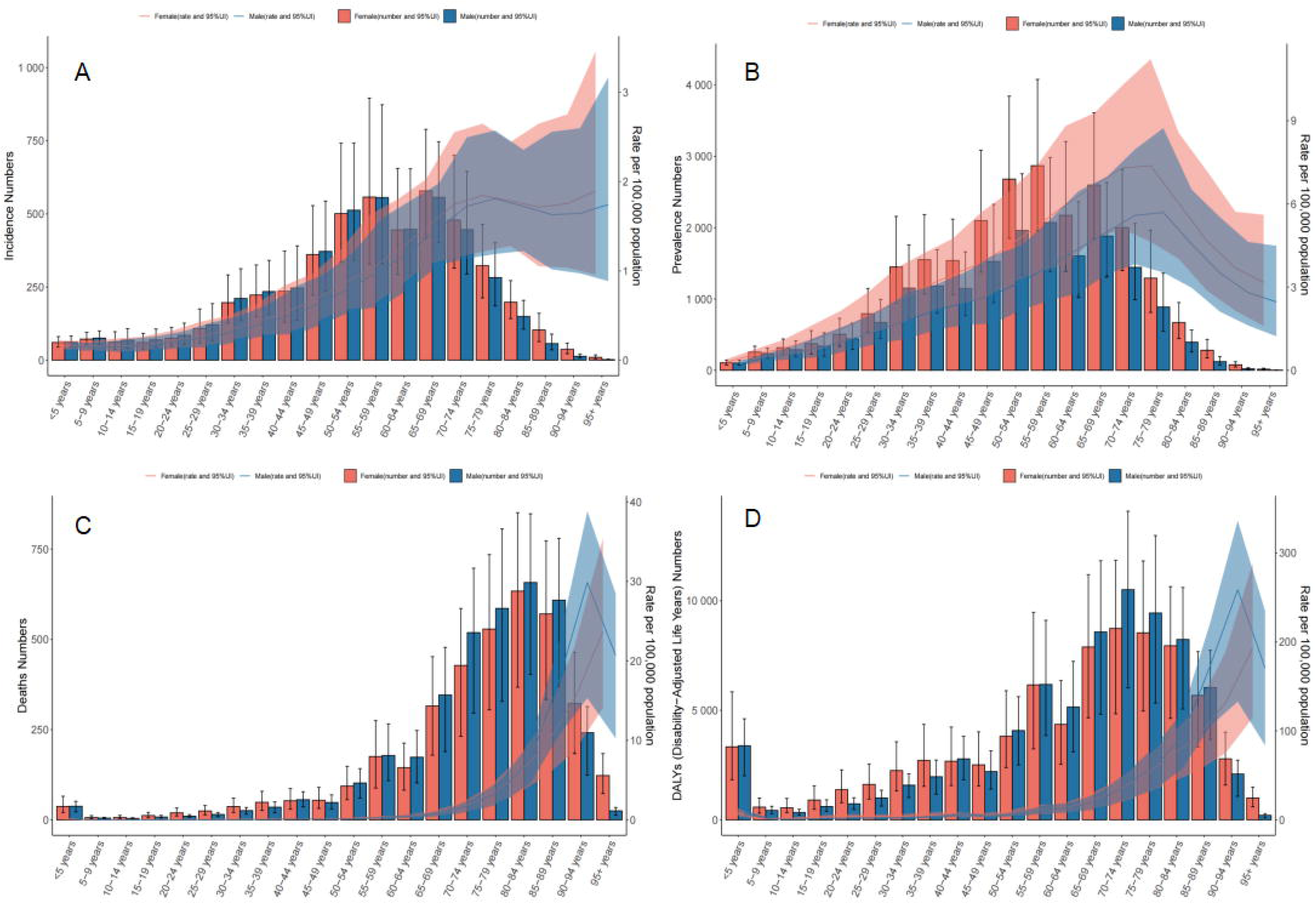
Illustrates the incidence, prevalence, mortality, and disability-adjusted life years (DAYLs) associated with PAH in China for the year 2021, segmented by age and sex. Detailing the incidence (Panel A), prevalence (Panel B), mortality (Panel C), and Disability-Adjusted Life Years (DALYs) (Panel D) associated with the condition. Shading represents the upper and lower limits of the 95% uncertainty intervals (95% UIs).

Gender-specific analysis revealed distinct trends in the burden of PAH. Among individuals younger than 50-54 years, the number of incident cases was higher in males than in females. However, starting from the 55–59 age group, the number of incident cases in females gradually surpassed that of males. The highest number of incident cases in females was observed in the 65-69 age group (579 cases), while the peak incidence rate was observed in the 95+ age group (1.90 per 100,000) (**Figure 1A, Supplementary Table S1**). Prevalence was consistently higher in females across all age groups. The largest number of prevalent cases in females was recorded in the 55-59 years age group (2869 cases), while the highest prevalence rate occurred in the 75-79 years age group (7.38/100,000) (**Figure 1B, Supplementary Table S2**). Mortality data showed that males aged 55-94 years had higher death cases compared to females in the same age range. Among females, the highest number of deaths occurred in the 80-84 age group (634 cases), while the highest death rate was observed in the 90-94 age group (29.86 per 100,000) (**Figure 1C, Supplementary Table S3**). For DAYLs, males had higher rates than females starting from the 50-54 age group, except in the 95+ age group. The DAYLs rate peaked at 258.70 per 100,1000 in males aged 90-94 years and at 193.40 per 100,000 in females aged 95+ years (**Figure 1D, Supplementary Table S4**).

### Trends of Pulmonary Arterial Hypertension from 1990 to 2021

From 1990 to 2021, the ASIR of PAH per 100,000 decreased slightly from 0.51 (95% UI: 0.41,0.62) to 0.50 (95% UI: 0.40,0.60), with an average annual percent change (EAPC) of −0.12 (95% CI: −0.17,-0.08). The decline in ASIR was more pronounced in females (−0.21, 95% CI: −0.26, −0.17) compared to males (−0.03, 95% CI: −0.09, 0.02) (**Table 1**). In contrast, the ASPR per 100,000 increased from 2.07(95%UI:1.68,2.54) in 1990 to 2.24(95%UI:1.81,2.75) in 2021, with an EAPC of 0.39(95% CI:0.34,0.44). The increase in ASPR was faster in males at 0.56(95%CI:0.51,0.62) than in females at 0.26(0.21,0.32) (**Table 2**). The ASDR per 100,000 decreased from 0.61 (95% UI: 0.46–0.83) in 1990 to 0.42 (95% UI: 0.28, 0.51) in 2021, with an EAPC of −0.61 (95% UI: −1.00 to, 0.22). The decline in ASDR was more pronounced in females (−0.87, 95% CI: −1.18,-0.57) compared to males (−0.37, 95% CI: −0.85, 0.12) (**Table 3**). Similarly, the age-standardized DALY rate per 100,000 declined significantly from 16.18(95%UI:12.63,21.60) in 1990 to 8.95(95%UI:6.04,11.13) in 2021 with an EAPC of −1.38(95%CI: −1.74,-1.01). The decline in DAYL rates was faster in females (−1.56,95%CI:-1.85,-1.27) compared to males (−1.17,95%CI:-1.62,-0.72) (**Table 4)**.

**Table 3.**
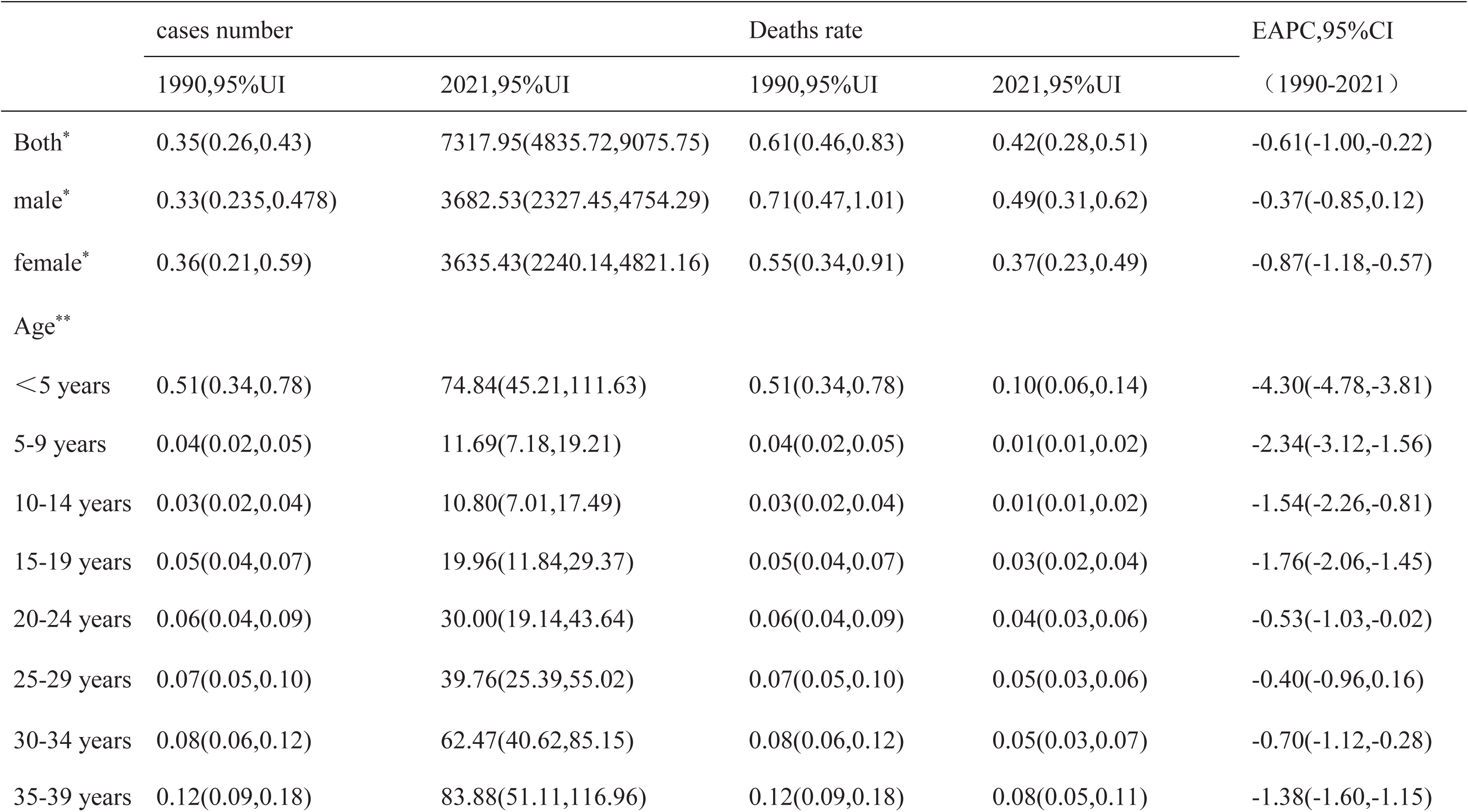
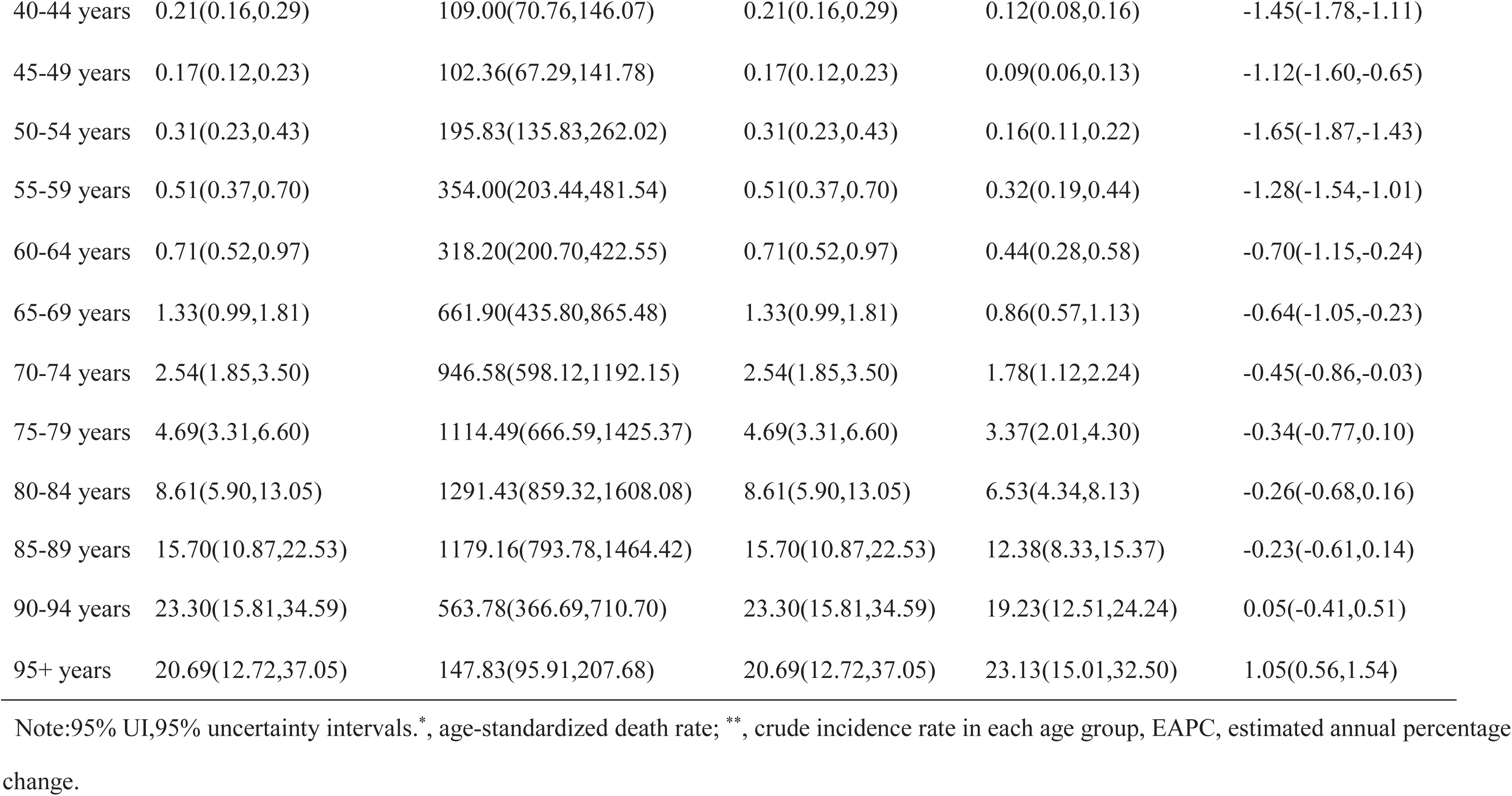
presents the number of cases and age-standardized death rates (ASDR) of pulmonary arterial hypertension in China for the years 1990 and 2021, along with their temporal trends from 1990 to 2021.

**Table 4.** presents the number of cases and age-standardized disability-adjusted life years of pulmonary arterial hypertension in China for the years 1990 and 2021, along with their temporal trends from 1990 to 2021.

|  | cases number |  | DAYLs rate |  | EAPC,95%CI<br>(1990-2021) |
| --- | --- | --- | --- | --- | --- |
|  | 1990,95%UI | 2021,95%UI | 1990,95%UI | 2021,95%UI |  |
| Both* | 12.73(9.85,17.22) | 150940.71(99583.31, 186503.40) | 16.18(12.63,21.60) | 8.95(6.04,11.13) | -1.38(-1.74,-1.01) |
| male* | 12.37(9.68,17.12) | 75517.87(48308.12,98062.18) | 17.09(12.91,23.32) | 9.48(6.05,12.04) | -1.17(-1.62,-0.72) |
| female* | 13.10(7.68,20.99) | 75422.84(47178.85,104591.34) | 15.80(9.38,25.61) | 8.78(5.66,12.69) | -1.56(-1.85,-1.27) |
| Age** |  |  |  |  |  |
| < 5 years | 45.87(30.23,69.64) | 6716.75(4072.71,9993.89) | 45.87(30.23,69.64) | 8.65(5.24,12.87) | -4.28(-4.77,-3.80) |
| 5-9 years | 2.93(2.00,4.25) | 1014.59(646.32,1633.79) | 2.93(2.00,4.25) | 1.06(0.68,1.71) | -2.27(-3.03,-1.50) |
| 10-14 years | 2.04(1.44,2.97) | 892.01(590.06,1407.19) | 2.04(1.44,2.97) | 1.04(0.69,1.63) | -1.46(-2.15,-0.76) |
| 15-19 years | 3.79(2.70,5.31) | 1512.99(930.11,2205.98) | 3.79(2.70,5.31) | 2.03(1.25,2.95) | -1.70(-2.00,-1.40) |
| 20-24 years | 4.06(2.91,6.06) | 2115.52(1378.67,3054.40) | 4.06(2.91,6.06) | 2.89(1.88,4.17) | -0.50(-0.99,-0.01) |
| 25-29 years | 4.44(3.12,6.42) | 2622.94(1723.80,3574.88) | 4.44(3.12,6.42) | 3.03(1.99,4.13) | -0.38(-0.92,0.17) |
| 30-34 years | 4.86(3.39,7.22) | 3837.34(2581.33,5136.88) | 4.86(3.39,7.22) | 3.17(2.13,4.24) | -0.65(-1.05,-0.25) |
| 35-39 years | 6.75(4.84,9.53) | 4684.11(2992.15,6411.03) | 6.75(4.84,9.53) | 4.42(2.82,6.05) | -1.30(-1.52,-1.09) |
| 40-44 years | 10.29(7.67,14.01) | 5457.32(3571.11,7250.93) | 10.29(7.67,14.01) | 5.96(3.90,7.92) | -1.40(-1.72,-1.08) |
| 45-49 years | 7.35(5.38,10.26) | 4717.28(3214.58,6438.44) | 7.35(5.38,10.26) | 4.28(2.91,5.84) | -1.05(-1.50,-0.59) |
| 50-54 years | 12.22(9.13,16.64) | 7890.60(5515.25,10361.60) | 12.22(9.13,16.64) | 6.53(4.56,8.57) | -1.57(-1.78,-1.36) |
| 55-59 years | 17.42(12.94,23.82) | 12334.77(7304.55,16688.40) | 17.42(12.94,23.82) | 11.22(6.64,15.18) | -1.23(-1.48,-0.97) |
| 60-64 years | 20.99(15.49,28.52) | 9507.08(6127.38,12496.60) | 20.99(15.49,28.52) | 13.02(8.39,17.12) | -0.67(-1.12,-0.22) |
| 65-69 years | 32.73(24.44,44.48) | 16458.81(10976.93,21324.74) | 32.73(24.44,44.48) | 21.46(14.31,27.80) | -0.62(-1.02,-0.21) |
| 70-74 years | 51.33(37.52,70.57) | 19235.97(12257.42,24197.57) | 51.33(37.52,70.57) | 36.09(23.00,45.40) | -0.44(-0.85,-0.03) |
| 75-79 years | 75.76(53.63,106.36) | 17970.33(10826.61,22948.90) | 75.76(53.63,106.36) | 54.26(32.69,69.29) | -0.36(-0.78,0.07) |
| 80-84 years | 108.39(74.33,164.02) | 16175.47(10777.87,20120.02) | 108.39(74.33,164.02) | 81.73(54.46,101.66) | -0.28(-0.69,0.14) |
| 85-89 years | 157.37(108.88,225.59) | 11712.77(7890.64,14535.96) | 157.37(108.88,225.59) | 122.96(82.84,152.60) | -0.25(-0.62,0.12) |
| 90-94 years | 201.89(137.18,299.51) | 4875.81(3172.21,6140.48) | 201.89(137.18,299.51) | 166.30(108.19,209.43) | 0.04(-0.42,0.50) |
| 95+ years | 171.31(105.31,306.59) | 1208.26(785.27,1693.75) | 171.31(105.31,306.59) | 189.06(122.87,265.02) | 1.01(0.52,1.50) |

Note:95% UI,95% uncertainty intervals.DAYLs,disability-adjusted life years;\*, age-standardized of rate; \*\*, crude incidence rate in each age group. EAPC, estimated annual percentage change.

### Temporal Trends of Pulmonary Arterial Hypertension Burdens in China

Using Joinpoint models, the age-standard incidence rate (ASIR) of PAH showed three declining phases followed by a significant increase. From 1990 to 2000, the ASIR declined at an annual percentage change (APC) of −0.444%, followed by a slower decline from 2000 to 2008 (APC = −0.050%) and a moderate decline from 2008 to 2016 (APC = −0.223%). However, from 2016 to 2021, the ASIR significantly increased with an APC of 0.712%. Over the entire study period, the average annual percent change (AAPC) was 0 **(Figure 2A).** The age-standard prevalence rate (ASPR) showed a steady increase from 1990 to 2013, with APCs of 0.287% (1990-1995) and 0.531% (1995-2013). From 2013 to 2019, the prevalence trend increased moderately (APC = 0.068%), but from 2019 to 2021, the ASPR declined significantly (APC = −1.471%). The overall AAPC for the study period was 0.005% (95%CI: 0.005% to 0.006%, *P*<0.05) (**Figure 2B**). The ASDR decreased significantly by −2.548% per year from 1990 to 2001, followed by a slower decline from 2001 to 2006 (APC = −0.303%). The death rate increased from 2006 to 2010 (APC = −6.684%), but declined rapidly from 2010 to 2021 (APC = −3.852%). The overall AAPC for the study period was −0.007% (95%CI: −0.008% to 0.007%, *P* > 0.05) (**Figure 2C**). By contrast, the age-standardized DAYL rate decreased significantly from 1990 to 2004 (APC= −3.096%), followed by an increase from 2004 to 2011(APC = 4.519%). From 2011 to 2021, the DAYL rate decreased again, with APCs of −4.867% (2011-2017) and −2.574% (2017-2021). The overall AAPC of DAYL for the study period was −0.257% (95%CI:-0.268% to 0.247%, *P*<0.05) (**Figure 2D**).

**Figure 2.**
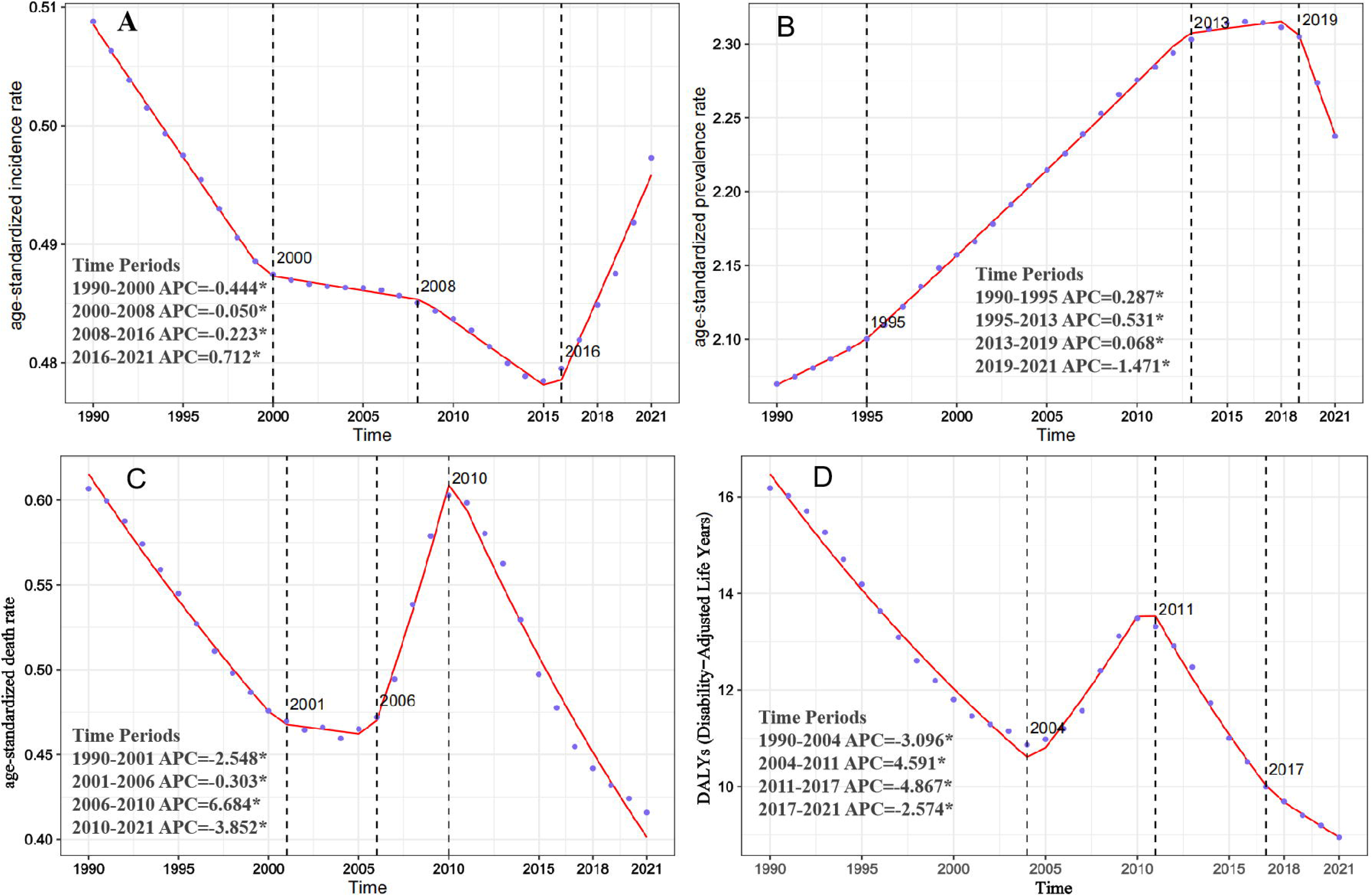
Average Annual Percentage Changes (AAPCs) in Pulmonary Arterial Hypertension Metrics in China (1990-2021).Panel A: Trends in age-standardized incidence rate; Panel B: Trends in age-standardized prevalence rate; Panel C: Trends in age-standardized mortality rate; Panel D: Trends in age-standardized disability-adjusted life years (DALYs). Abbreviations: ASIR, age-standardized incidence rates; ASPR, age-standardized prevalence rates; ASDR, age-standardized death rates;ASYR, age-standardized disability-adjusted life year; APC, annual percent change.

### Age, Period, and Cohort Effects on Incidence and Death Rates

#### Age Effect

The age-period-cohort (APC) analysis revealed distinct PAH incidence and mortality patterns. After controlling for period and cohort effects, the age effect showed that incidence rate risk substantially increased with age, reaching a peak in the 75-79 age group, followed by a slight decrease before rising again in the 95+ age group (**Figure 3A, Supplementary Table S5)**. From age groups< 5 years to 75-79 years, the incidence increased by 10.81-fold. The PAH death rate showed a continuous increase with advancing age from the 70-74 age group, with the death rate in the 95+ age group being approximately 38.81 times higher than in the <5 age group (**Figure 3D, Supplementary Table S5**).

**Figure 3:**
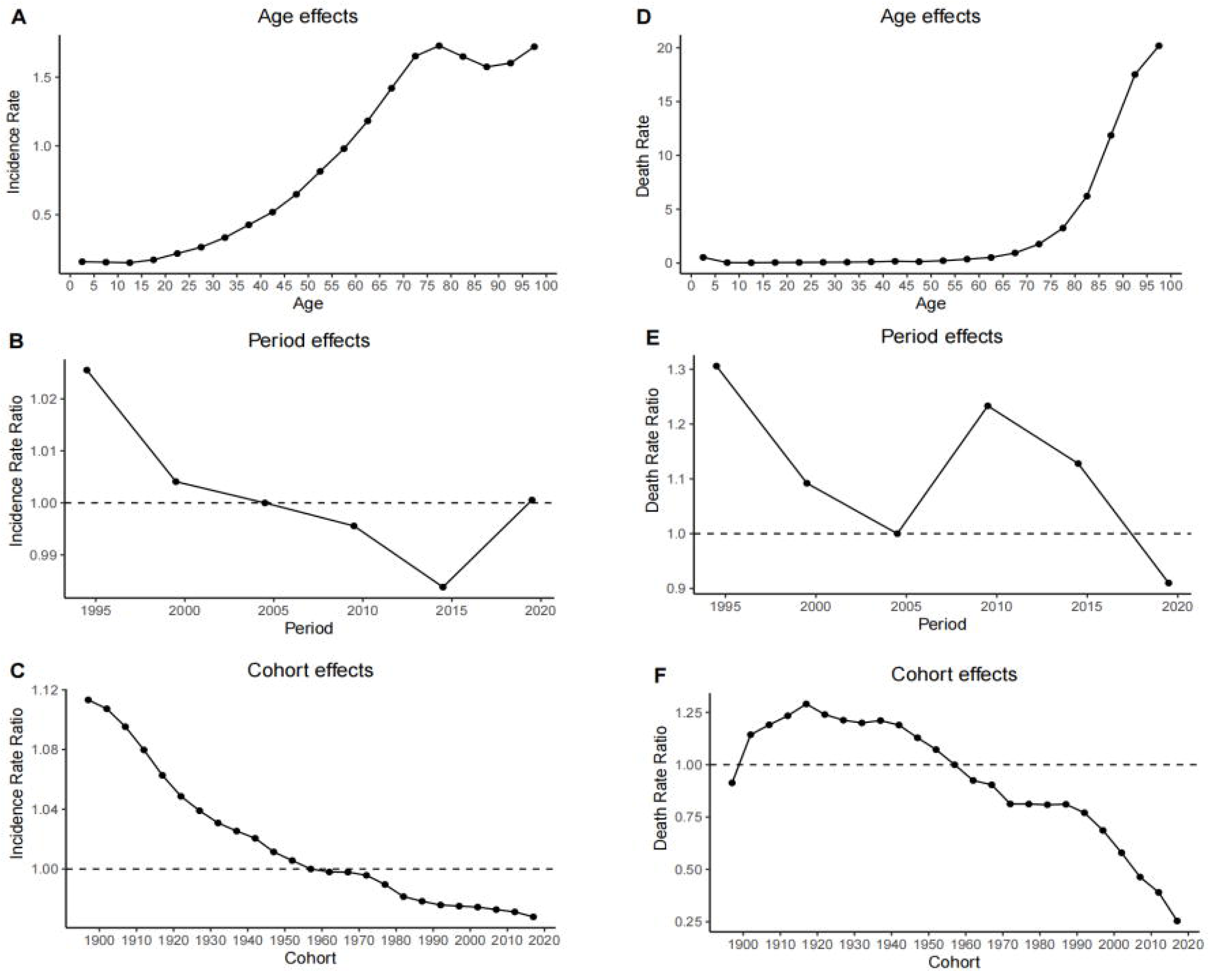
Age, Period, and Cohort Effects on Pulmonary Arterial Hypertension Incidence and Mortality in China: Relative Risk Analysis.Panel A: Age effects on incidence relative risk; Panel B: Period effects on incidence relative risk; Panel C: Cohort effects on incidence relative risk; Panel D: Age effects on mortality relative risk;Panel E: Period effects on mortality relative risk; Panel F: Cohort effects on mortality relative risk.

#### Period Effect

The period effect analysis revealed that incidence risk gradually declined from 1995 (RR = 1.03, 95% CI: 0.99 to 1.07) to 2015 (RR = 0.98, 95% CI: 0.95 to 1.02), followed by a slight increase from 2015 to 2020 (RR = 1.00, 95% CI: 0.96 to 1.04) **(Figure 3B, Supplementary Table S5).** Mortality rates showed a decline from 1995 (RR = 1.31, 95% CI: 1.25 to 1.37) to 2005 (RR = 1.09, 95% CI: 1.04 to 1.14), followed by an increase from 2005 to 2010 (RR = 1.23, 95% CI: 1.18 to 1.28), and then another decline from 2010 to 2020 (RR = 0.91, 95% CI: 0.87 to 0.95) **(Figure 3E, Supplementary Table S5).**

#### Cohort Effect

The cohort effects analysis of incidence risk (**Figure 3C, Supplementary Table S5**) revealed an inverse relationship between birth cohort and risk levels. The 1895-1899 cohort exhibited the highest incidence risk (*RR*=1.11, 95% CI: 0.14-8.13). A distinct pattern emerged: cohorts from 1895-1955 showed elevated incidence rate ratios (*RR* >1), while cohorts from 1981-2021 demonstrated reduced ratios (*RR* <1). Overall, the incidence rate ratio displayed a declining trend. For mortality risk, the cohort effects analysis (**Figure 3F, Supplementary Table S5**) showed that cohorts born between 1895-1919 experienced elevated death rate ratios (*RR* >1), peaking in the 1915-1919 cohort (*RR*=1.29, 95% CI: 1.18-1.41). From 1920 onward, death rate ratios steadily declined, with the most pronounced decrease observed in cohorts 1991-2021.

### Projected Burden of Pulmonary Arterial Hypertension from 2021 to 2050

Using data from the GBD database (1990-2021), we employed the Bayesian Age-Period-Cohort (BAPC) model to project the future burden of PAH in China through 2050. The ASIR for China’s total population is projected to decline modestly from 0.493 per 100,000 in 2021 to 0.486 per 100,000 by 2050, representing a 1.42% decrease **(Figure 4A)**. Regarding gender-specific ASIR, the projections showing an increase in male from 0.488 to 0.493 per 100,000 and a decrease in female from 0.493 to 0.475 per 100,000 **(Supplementary Table S6)**. In contrast, the ASPR is expected to reach 2.443 per 100,000 by 2050, representing an 8.29% increase from 2021 levels **(Figure 4B)**, with male rising from 1.907 to 2.163 per 100,000 and female increasing from 2.573 to 2.725 per 100,000, respectively **(Supplementary Table S6)**. The ASDR shows a substantial projected decline for the total population, decreasing from 0.418 per 100,000 to 0.236 per 100,000 by 2050, representing a 43.54% reduction **(Figure 4C)**, with male decreasing from 0.498 to 0.283 per 100,000 and female declining from 0.374 to 0.211 per 100,000 **(Supplementary Table S6)**. The age-standardized DALY rate is projected to demonstrate the most substantial improvement, with a remarkable decrease of 49.69%, reaching 4.370 per 100,000 by 2050 (**Figure 4D**)/ Specfically, the rate for males is expected to decline to 4.942 per 100,000, while the rate for female will decrease to 4.067 per 100,000 (**Supplementary Table S6**).

**Figure 4.**
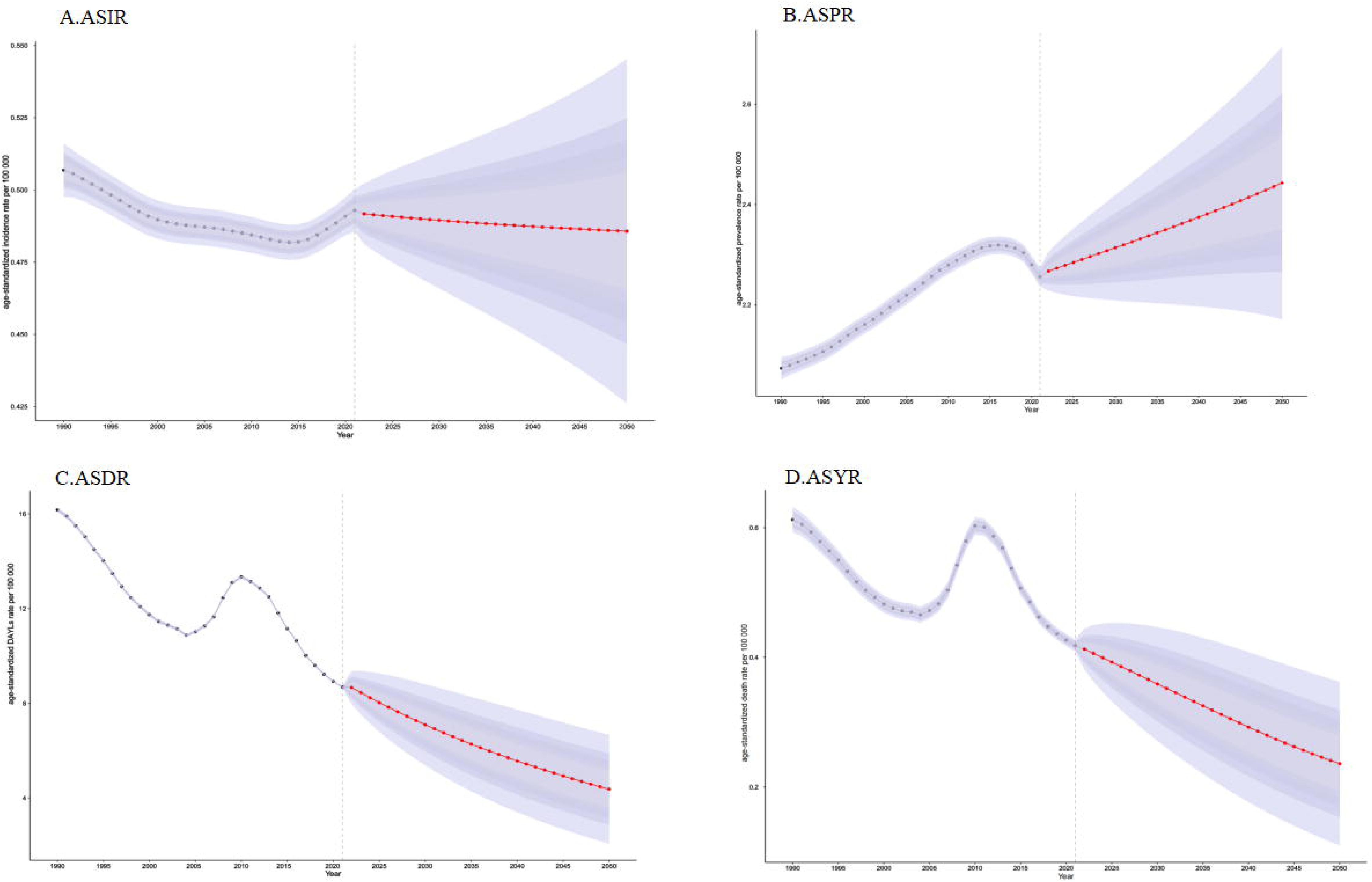
Temporal Trends and Projections of Pulmonary Arterial Hypertension Burden in China, 1990-2050.Panel A: Projected age-standardized incidence rate; Panel B: Projected age-standardized prevalence rate; Panel C: Projected age-standardized mortality rate; Panel D: Projected age-standardized disability-adjusted life years (DALY). Red dot lines and blue shaded regions represent the predicted trend and 95% CI. Abbreviations: ASIR, age-standardized incidence rates;ASPR:age-standardized prevalence rates;ASDR:age-standardized death rates; ASYR,age-standardized disability-adjusted life year.CI, confidence interval.

## Discussion

This study represents the first comprehensive analysis of PAH disease burden in China based on the GBD 2021 data, providing crucial epidemiological insights from 1990 to 2021. Our findings reveal that in 2021, the burden of PAH in China experienced an incidence rate of 0.51 per 100,000 population, a prevalence rate of 2.24 per 100,000, a mortality rate of 0.42 per 100,000, and a DALY rate of 8.95 per 100,000. Compared to 1990, while the incidence, mortality, and DALYs have declined, the prevalence has increased. The observed prevalence increase might be attributed to several factors: improved early diagnostic techniques, an aging population, increased exposure to risk factors (including tobacco smoke, drug abuse, and environmental pollutants), and lacking awareness among healthcare providers and the public^4,37–42^. Our findings align with previous registry-based studies. The French registry (2002-2003) reported an incidence of 0.24 and a prevalence of 1.5 per 100,000^15^, while the US REVEAL registry (2006-2007) documented an incidence of 0.20 and a prevalence of 1.06 per 100,000^43^. The current global GBD estimate of 2.28 prevalent cases per 100,000 in 2021 closely matches our finding of 2.24 per 100,000 in China^21^, validating the reliability of GBD data for PAH surveillance. Nevertherless, the mortality rate of PAH in China (0.42 per 100,000) exceeds the average death rate globally (0.27 per 100,000). This disparity likely stems from unbalanced medical resource distribution and delayed diagnosis, particularly in rural areas, leading to more advanced-stage diagnoses when treatment efficacy is reduced^27,44–46^. Therefore, in the future, efforts should focus on improving access to medical resources and early diagnosis in rural areas.

Gender-specific analysis reveals higher incidence (0.50 vs 0.49 per 100,000) and prevalence (2.57 vs 1.97 per 100,000) in females compared to males. However, males experience higher mortality (0.49 vs 0.37 per 100,000) and DALY rates (9.48 vs 8.78 per 100,000). These discrepancies align with findings from previous studies, which indicate that while females are more susceptible to developing PAH, they tend to exhibit better right ventricular (RV) function and higher survival rates compared to males^21,47–49^. This phenomenon, often referred to as the "estrogen paradox" or "estrogen puzzle," is partly attributed to the protective effects of estrogen on vascular and RV function^50,51^. Additionally, females are more likely to respond positively to treatment with prostacyclin analogs, which may contribute to their lower mortality rates. These factors affirm the long-standing recognition that PAH is more prevalent in females but often more severe in males^52^.

Historically, PAH was believed to have a higher incidence among younger individuals, particularly younger females^14,49,53,54^. This view arose from studies focused on specific types of PAH, which are more common in younger populations. However, as research has advanced, it has become evident that PAH incidence increases significantly in older adults. Our study shows that the peak age-standardized incidence and prevalence occur in the 75–79 age group, while mortality rates increase with advancing age. These findings are consistent with global GBD estimates^21^, suggesting that the growing recognition of PAH in older populations may be due to the availability of PAH-specific therapies and the widespread use of Doppler echocardiography^48^, improving the diagnosis in older adults.

Looking ahead, our 30-year projections suggest declining overall incidence and mortality. This trend is likely driven by advancements in PAH management, including the use of endothelin receptor antagonists (ERAs), phosphodiesterase-5 inhibitors (PDE5is), prostacyclin analogs, and novel therapeutic strategies such as initial combination therapy, exercise training, and psychological support^2,19,55–57^. These treatments have demonstrably improved symptoms, slowed disease progression, and extended survival, contributing to reduced mortality and morbidity. However, the prevalence rate is projected to rise, which might be driven by the aging population, unhealthy lifestyle factors (e.g., smoking, obesity, and physical inactivity), and environmental pollution^58–61^. Additionally, advancements in diagnostic tools, including echocardiography, CT, MRI, hemodynamic variables (e.g., right atrial pressure, cardiac index, and venous oxygen saturation), and biomarker utilization could also increase the number of diagnosed cases^2,62–66^. This increase should not be interpreted as a genuine rise in incidence but rather as a result of improved diagnostic accuracy. These findings underscore the need for continued surveillance, targeted interventions, and further research to address the complex epidemiological trends of PAH in China. Future studies should focus on quantifying the impact of various risk factors and developing effective public health strategies to manage this challenging condition.

This study has several limitations. First, it focuses exclusively on group 1 PAH and does not include groups 2 – 5 pulmonary hypertension, which limits the scope of our findings. Second, the diagnosis of PAH is complex, and many primary medical institutions may lack the necessary diagnostic equipment and trained professionals. This can lead to a high prevalence of missed or misdiagnosed cases, potentially resulting in an underestimation of PAH cases in the GBD database. Third, we could not obtain data on pulmonary hypertension across different provinces in China, preventing us from analyzing regional and urban-rural disparities in the burden of pulmonary hypertension. Future research should prioritize large-scale, high-quality epidemiological studies that integrate multiple data sources to more accurately assess the epidemiological characteristics of PAH in China. Furthermore, improving PAH diagnostic and reporting systems is crucial for enhancing data quality. These advancements will facilitate better utilization of the GBD database and other resources, enabling a more precise evaluation of the PAH disease burden in China. However, despite these limitaitons, this study still could offer valuable insights into the temporal trends and overall burden of PAH in China, providing an important foundation for future research and public health planning.

## Conclusion

PAH remains a significant public health challenge in China, with an estimated 41,135 individuals affected in 2021, leading to 7,318 deaths and 150,941 DALYs. The burden is particularly pronounced among women and older adults, underscoring the need for gender- and age-specific strategies to address this condition. While projections indicate a continued decline in PAH incidence and mortality rates, the prevalence of PAH is expected to rise, driven by demographic shifts and persistent risk factors. These findings emphasize the urgent need for sustained efforts to enhance early detection, expand access to specialized care, and implement targeted public health interventions to mitigate the growing burden of PAH in China. Future research should prioritize addressing regional disparities, quantifying the influence of risk factors, and developing innovative strategies to reduce the disease burden and improve patient outcomes.

## Data Availability

All data produced are available online at http://ghdx.healthdata.org/gbd-results-tool

## Contributors

LZ conceptualised the study. SSW completed the first draft of the manuscript. ML,YHH and HLW contributed to the study methodology. YSC and ZWL was involved in the interpretation of the data. YSC, JG and LZ provided critical comments on drafts of the manuscript. LZ was responsible for the decision to submit the manuscript. All authors participated in the review of the manuscript and read and approved the final manuscript. All authors had access to the data in the study and had final responsibility to submit for publication.

## Data sharing statement

The data from this study can be accessed openly through the GBD 2021 online database, as outlined in the Methods section.

## Acknowledgments

We acknowledge the exceptional contributions made by the collaborators of the Global Burden of Diseases, Injuries, and Risk Factors Study 2021. We sincerely appreciate the IHME institution for providing the GBD data.

