## Supplemental document for "Epidemiological Burden and Projections of Pulmonary Arterial Hypertension in China: An Analysis from the Global Burden of Disease Study 2021"

**Table S1. Numbers and Crude Incidence Rates of Pulmonary Arterial Hypertension by Age and Sex in China, 2021**

|  | **Females** | | **male** | |
| --- | --- | --- | --- | --- |
| **Age group** | **incidence cases** | **Incidence rate** | **incidence cases** | **Incidence rate** |
| ＜5 years | 60.42(44.72,80.56) | 0.17(0.12,0.22) | 61.34(44.96,82.97) | 0.15(0.11,0.20) |
| 5-9 years | 72.17(52.96,95.00) | 0.16(0.12,0.21) | 72.17(52.96,95.00) | 0.16(0.12,0.21) |
| 10-14 years | 61.99(36.68,97.28) | 0.15(0.09,0.24) | 68.85(39.99,108.10) | 0.15(0.09,0.24) |
| 15-19 years | 60.20(38.02,91.63) | 0.17(0.11,0.26) | 69.66(43.98,106.62) | 0.17(0.11,0.27) |
| 20-24 years | 75.33(47.74,112.13) | 0.22(0.14,0.33) | 85.35(54.10,126.49) | 0.22(0.14,0.33) |
| 25-29 years | 108.50(59.09,174.86) | 0.27(0.15,0.43) | 121.42(66.74,193.31) | 0.27(0.15,0.42) |
| 30-34 years | 196.81(126.27,291.35) | 0.34(0.22,0.50) | 211.30(134.96,312.62) | 0.34(0.22,0.50) |
| 35-39 years | 222.77(147.41,325.49) | 0.432(0.29,0.63) | 234.53(155.38,341.20) | 0.43(0.29,0.63) |
| 40-44 years | 235.47(129.11,373.12) | 0.53(0.29,0.84) | 246.99(137.00,390.77) | 0.53(0.29,0.83) |
| 45-49 years | 361.00(222.53,528.28) | 0.67(0.41,0.97) | 371.87(234.45,543.56) | 0.66(0.42,0.97) |
| 50-54 years | 501.21(334.37,741.78) | 0.84(0.56,1.24) | 512.48(341.13,742.30) | 0.84(0.56,1.21) |
| 55-59 years | 557.47(328.24,895.62) | 1.01(0.60,1.63) | 555.69(329.73,872.93) | 1.01(0.60,1.59) |
| 60-64 years | 445.25(292.99,655.43) | 1.22(0.81,1.80) | 447.43(295.89,654.18) | 1.22(0.81,1.79) |
| 65-69 years | 578.58(416.82,788.89) | 1.49(1.07,2.03) | 556.33(401.06,746.19) | 1.47(1.06,1.98) |
| 70-74 years | 478.86(314.17,700.38) | 1.75(1.15,2.55) | 446.14(294.32,645.34) | 1.73(1.14,2.50) |
| 75-79 years | 323.25(212.97,464.07) | 1.85(1.22,2.65) | 282.31(185.53,401.47) | 1.81(1.19,2.57) |
| 80-84 years | 198.28(142.15,271.76) | 1.78(1.28,2.45) | 149.54(106.02,204.35) | 1.72(1.22,2.36) |
| 85-89 years | 103.64(63.55,160.26) | 1.72(1.05,2.65) | 56.75(35.41,89.03) | 1.63(1.02,2.56) |
| 90-94 years | 37.25(22.01,58.43) | 1.75(1.04,2.75) | 13.29(7.86,21.02) | 1.64(0.97,2.60) |
| 95+ years | 9.83(4.96,17.92) | 1.90(0.96,3.46) | 2.11(1.07,3.83) | 1.74(0.89,3.17) |

**Table S2. Numbers and Crude Prevalence Rates of Pulmonary Arterial Hypertension by Age and Sex in China, 2021**

| **Age group** | **Females** | | **male** | |
| --- | --- | --- | --- | --- |
|  | **prevalence cases** | **prevalence rate** | **prevalence cases** | **prevalence rate** |
| ＜5 years | 106.75(76.20,144.90) | 0.30(0.21,0.40) | 101.72(71.70,140.40) | 0.24(0.17,0.34) |
| 5-9 years | 257.17(186.03,338.18) | 0.57(0.42,0.75) | 236.23(169.34,309.66) | 0.46(0.33,0.61) |
| 10-14 years | 317.25(217.50,439.26) | 0.79(0.541,1.09) | 288.90(189.83,411.85) | 0.63(0.41,0.90) |
| 15-19 years | 373.59(226.37,556.75) | 1.08(0.65,1.61) | 339.34(196.53,526.49) | 0.85(0.49,1.31) |
| 20-24 years | 504.82(334.55,728.13) | 1.47(0.98,2.12) | 441.15(293.44,661.94) | 1.14(0.76,1.70) |
| 25-29 years | 793.65(531.03,1147.05) | 1.94(1.30,2.81) | 670.81(445.98,993.32) | 1.47(0.98,2.18) |
| 30-34 years | 1449.84(943.69,2157.48) | 2.48(1.61,3.69) | 1153.19(726.56,1759.47) | 1.84(1.16,2.81) |
| 35-39 years | 1551.80(1069.69,2182.56) | 3.01(2.07,4.23) | 1182.85(800.31,1690.38) | 2.18(1.47,3.11) |
| 40-44 years | 1539.46(1053.80,2118.75) | 3.45(2.36,4.75) | 1145.84(766.28,1660.67) | 2.44(1.63,3.54) |
| 45-49 years | 2098.70(1374.89,3085.80) | 3.87(2.53,5.69) | 1526.09(948.02,2327.17) | 2.72(1.69,4.15) |
| 50-54 years | 2680.93(1850.84,3843.35) | 4.49(3.10,6.44) | 1961.24(1338.96,2754.97) | 3.21(2.19,4.51) |
| 55-59 years | 2869.10(1953.11,4078.10) | 5.21(3.55,7.41) | 2072.79(1385.91,2984.37) | 3.78(2.53,5.44) |
| 60-64 years | 2172.76(1387.17,3207.52) | 5.97(3.81,8.82) | 1604.67(1021.22,2365.73) | 4.38(2.79,6.46) |
| 65-69 years | 2597.62(1841.25,3611.05) | 6.67(4.73,9.27) | 1882.89(1308.61,2632.33) | 4.99(3.47,6.98) |
| 70-74 years | 2001.99(1397.07,2818.30) | 7.30(5.09,10.27) | 1443.88(989.36,2059.93) | 5.58(3.83,7.97) |
| 75-79 years | 1292.46(811.82,1964.90) | 7.38(4.64,11.22) | 887.83(550.20,1362.03) | 5.69(3.53,8.73) |
| 80-84 years | 668.96(446.31,950.69) | 6.02(4.02,8.55) | 394.69(258.26,565.41) | 4.55(2.98,6.52) |
| 85-89 years | 281.54(173.35,433.72) | 4.66(2.87,7.18) | 122.32(73.68,192.30) | 3.51(2.12,5.52) |
| 90-94 years | 78.30(45.56,121.42) | 3.69(2.15,5.72) | 22.68(13.07,37.43) | 2.80(1.62,4.63) |
| 95+ years | 16.43(8.29,29.04) | 3.17(1.60,5.61) | 2.98(1.48,5.43) | 2.47(1.23,4.49) |

**Table S3. Numbers and Crude Death Rates of Pulmonary Arterial Hypertension by Age and Sex in China, 2021**

| **Age group** | **Females** | | | **male** | |
| --- | --- | --- | --- | --- | --- |
|  | **death cases** | **death rate** | | **death cases** | **death rate** |
| ＜5 years | 37.19(20.42,65.36) | 0.10(0.06,0.18) | 37.66(22.39,51.38) | | 0.09(0.05,0.12) |
| 5-9 years | 6.74(3.41,11.74) | 0.02(0.01,0.03) | 4.95(3.13,7.17) | | 0.01(0.01,0.01) |
| 10-14 years | 6.77(3.92,12.23) | 0.02(0.01,0.03) | 4.03(2.48,5.98) | | 0.01(0.01,0.01) |
| 15-19 years | 12.00(5.99,20.84) | 0.04(0.02,0.06) | 7.96(4.94,12.23) | | 0.02(0.01,0.03) |
| 20-24 years | 19.73(10.95,32.97) | 0.06(0.03,0.10) | 10.27(6.44,14.44) | | 0.03(0.02,0.04) |
| 25-29 years | 24.71(13.86,39.72) | 0.06(0.03,0.10) | 15.05(9.06,20.48) | | 0.03(0.02,0.05) |
| 30-34 years | 36.81(20.52,60.22) | 0.06(0.04,0.10) | 25.67(15.80,34.45) | | 0.04(0.03,0.06) |
| 35-39 years | 48.68(25.89,79.14) | 0.09(0.05,0.15) | 35.19(20.05,49.69) | | 0.07(0.04,0.09) |
| 40-44 years | 53.04(29.39,86.62) | 0.12(0.07,0.19) | 55.96(36.04,77.77) | | 0.12(0.08,0.17) |
| 45-49 years | 54.14(31.83,89.86) | 0.10(0.06,0.17) | 48.22(29.95,69.32) | | 0.09(0.05,0.12) |
| 50-54 years | 93.71(55.90,148.46) | 0.16(0.09,0.25) | 102.12(60.15,141.97) | | 0.17(0.10,0.23) |
| 55-59 years | 175.64(88.26,275.01) | 0.32(0.16,0.50) | 178.36(108.95,266.16) | | 0.33(0.20,0.49) |
| 60-64 years | 144.52(82.28,212.77) | 0.40(0.23,0.59) | 173.68(102.58,247.71) | | 0.47(0.28,0.68) |
| 65-69 years | 315.83(179.258,452.15) | 0.81(0.46,1.16) | 346.07(189.36,477.94) | | 0.92(0.50,1.27) |
| 70-74 years | 427.81(231.24,585.68) | 1.56(0.84,2.13) | 518.77(295.78,697.20) | | 2.01(1.14,2.70) |
| 75-79 years | 528.60(304.91,734.96) | 3.02(1.74,4.20) | 585.89(328.12,805.80) | | 3.76(2.10,5.17) |
| 80-84 years | 633.85(367.61,850.88) | 5.70(3.31,7.65) | 657.58(403.02,847.88) | | 7.58(4.65,9.77) |
| 85-89 years | 570.58(333.83,773.01) | 9.44(5.52,12.79) | 608.59(369.27,779.79) | | 17.48(10.61,22.40) |
| 90-94 years | 241.51(124.01,314.04) | 29.86(15.33,38.83) | 322.26(183.51,463.64) | | 15.18(8.64,21.84) |
| 95+ years | 122.84(72.89,183.64) | 23.71(14.07,35.44) | 25.00(12.38,34.41) | | 20.67(10.23,28.45) |

**Table S4.Numbers and Crude DALY Rates of Pulmonary Arterial Hypertension by Age and Sex in China, 2021**

| **Age group** | **Females** | | **male** | |
| --- | --- | --- | --- | --- |
|  | **DAYLs cases** | **DAYLs rate** | **DAYLs cases** | **DAYLs rate** |
| ＜5 years | 3334.76(1835.74,5843.20) | 9.25(5.094,16.21) | 3381.99(2014.81,4611.66) | 8.124(4.840,11.078) |
| 5-9 years | 582.79(306.77,995.46) | 1.30(0.68,2.22) | 431.80(278.27,620.35) | 0.848(0.546,1.218) |
| 10-14 years | 553.37(334.69,978.67) | 1.38(0.833,2.434) | 338.65(214.29,486.01) | 0.736(0.466,1.057) |
| 15-19 years | 904.71(482.45,1550.35) | 2.62(1.40,4.48） | 608.28(391.11,918.14) | 1.518(0.976,2.291) |
| 20-24 years | 1380.85(782.71,2283.37) | 4.03(2.28,6.66) | 734.68(477.96,1010.22) | 1.890(1.230,2.599) |
| 25-29 years | 1619.30(949.02,2548.89) | 3.96(2.32,6.24) | 1003.64(628.77,1352.42) | 2.200(1.378,2.965) |
| 30-34 years | 2252.54(1317.33,3564.00) | 3.85(2.25,6.10) | 1584.80(1020.49,2099.18) | 2.528(1.628,3.349) |
| 35-39 years | 2715.01(1526.81,4357.28) | 5.26(2.96,8.45) | 1969.11(1170.49,2729.41) | 3.622(2.153,5.020) |
| 40-44 years | 2676.04(1567.29,4234.57) | 6.00(3.51,9.49) | 2781.28(1832.42,3815.44) | 5.927(3.905,8.131) |
| 45-49 years | 2510.89(1543.99,4015.20) | 4.63(2.85,7.40) | 2206.39(1402.59,3147.92) | 3.936(2.502,5.615) |
| 50-54 years | 3815.90(2384.24,5890.51) | 6.39(3.99,9.86) | 4074.70(2509.20,5621.57) | 6.664(4.104,9.194) |
| 55-59 years | 6157.18(3237.26,9467.91) | 11.18(5.88,17.20) | 6177.59(3850.55,9101.36) | 11.256(7.016,16.583) |
| 60-64 years | 4358.18(2533.20,6356.09) | 11.98(6.96,17.47） | 5148.891(3105.638,7230.190) | 14.058(8.479,19.741) |
| 65-69 years | 7888.44(4655.84,11186.89) | 20.25(11.949,28.71) | 8570.37(4818.19,11820.57) | 2.709(12.767,31.321) |
| 70-74 years | 8734.82(4838.94,11843.18) | 31.83(17.64,43.16) | 10501.15(6025.05,14079.86) | 40.613(23.302,54.454) |
| 75-79 years | 8532.64(4967.13,11811.76) | 48.71(28.36,67.43) | 9437.69(5333.27,12970.86) | 60.489(34.182,83.134) |
| 80-84 years | 7945.06(4628.86,10640.13) | 71.47(41.64,95.71) | 8230.41(5057.48,10600.58) | 94.875(58.300,122.197) |
| 85-89 years | 5674.92(3331.20,7674.22) | 93.88(55.11,126.96) | 6037.85(3668.76,7730.14) | 173.456(105.396,222.072) |
| 90-94 years | 2783.37(1588.53,3999.43) | 131.10(74.82,188.37) | 2092.44(1075.05,2720.12) | 258.698(132.914,336.302) |
| 95+ years | 1002.09(595.80,1494.98) | 193.40(114.98,288.52) | 206.17(101.01,283.99) | 170.467(83.516,234.808) |

Note：DAYLs,disability-adjusted life years.

**Table S5. Age-Period-Cohort (APC) Model Analysis of Pulmonary Arterial Hypertension Incidence and Mortality Rates in China**

| **Variables** | **incidence(95%CI)** | **death rate(95%CI)** |
| --- | --- | --- |
| **Age** |  |  |
| ＜5 years | 0.16(0.14,0.18) | 0.52(0.43,0.63) |
| 5-9 years | 0.16(0.14,0.17) | 0.03(0.03,0.04) |
| 10-14 years | 0.15(0.14,0.17) | 0.03(0.02,0.03) |
| 15-19 years | 0.17(0.16,0.19) | 0.05(0.04,0.06) |
| 20-24 years | 0.22(0.20,0.24) | 0.06(0.05,0.07) |
| 25-29 years | 0.26(0.25,0.28) | 0.07(0.06,0.08) |
| 30-34 years | 0.33(0.31,0.36) | 0.07(0.07,0.08) |
| 35-39 years | 0.43(0.40,0.45) | 0.11(0.09,0.12) |
| 40-44 years | 0.52(0.49,0.55) | 0.16(0.15,0.18) |
| 45-49 years | 0.65(0.62,0.68) | 0.13(0.11,0.14) |
| 50-54 years | 0.82(0.78,0.85) | 0.21(0.20,0.23) |
| 55-59 years | 0.98(0.94,1.02) | 0.35(0.33,0.38) |
| 60-64 years | 1.18(1.13,1.24) | 0.51(0.48,0.55) |
| 65-69 years | 1.42(1.35,1.50) | 0.93(0.86,1.01) |
| 70-74 years | 1.65(1.56,1.75) | 1.76(1.62,1.90) |
| 75-79 years | 1.73(1.62,1.84) | 3.25(2.99,3.52) |
| 80-84 years | 1.65(1.52,1.78) | 6.22(5.72,6.75) |
| 85-89 years | 1.57(1.41,1.76) | 11.86(10.89,12.92) |
| 90-94 years | 1.60(1.32,1.94) | 17.52(15.92,19.28) |
| 95+ years | 1.72(1.13,2.63) | 20.18(17.53,23.23) |
| **Period** |  |  |
| 1995 | 1.03(0.99,1.07) | 1.31(1.25,1.37) |
| 2000 | 1(0.97,1.04) | 1.09(1.04,1.14) |
| 2005 | 1(1,1) | 1(1,1) |
| 2010 | 1(0.96,1.03) | 1.23(1.18,1.28) |
| 2015 | 0.98(0.95,1.02) | 1.13(1.08,1.18) |
| 2020 | 1(0.96,1.04) | 0.91(0.87,0.95) |
| **Cohort** |  |  |
| 1895-1899 | 1.11(0.14,8.73) | 0.91(0.48,1.75) |
| 1900-1904 | 1.11(0.56,2.17) | 1.14(0.93,1.41) |
| 1905-1909 | 1.10(0.81,1.47) | 1.19(1.05,1.35) |
| 1910-1914 | 1.08(0.92,1.27) | 1.23(1.12,1.36) |
| 1915-1919 | 1.06(0.95,1.19) | 1.29(1.18,1.41) |
| 1920-1924 | 1.05(0.96,1.14) | 1.24(1.14,1.35) |
| 1925-1929 | 1.04(0.97,1.11) | 1.21(1.12,1.32) |
| 1930-1934 | 1.03(0.97,1.11) | 1.20(1.11,1.30) |
| 1935-1939 | 1.03(0.97,1.08) | 1.21(1.12,1.31) |
| 1940-1944 | 1.02(0.97,1.08) | 1.19(1.10,1.29) |
| 1945-1950 | 1.01(0.96,1.06) | 1.13(1.04,1.22) |
| 1951-1955 | 1.01(0.96,1.05) | 1.07(0.99,1.16) |
| 1956-1960 | 1(1,1) | 1(1,1) |
| 1961-1965 | 1(0.95,1.05) | 0.92(0.85,1.01) |
| 1966-1970 | 1(0.95,1.05) | 0.90(0.82,1.00) |
| 1971-1975 | 1(0.94,1.05) | 0.81(0.73,0.91) |
| 1976-1980 | 1(0.93,1.06) | 0.81(0.71,0.93) |
| 1981-1985 | 0.98(0.91,1.06) | 0.81(0.69,0.95) |
| 1986-1990 | 0.98(0.91,1.06) | 0.81(0.69,0.96) |
| 1991-1995 | 0.98(0.89,1.07) | 0.77(0.64,0.93) |
| 1996-2000 | 0.98(0.88,1.09) | 0.69(0.56,0.84) |
| 2001-2005 | 0.97(0.86,1.10) | 0.58(0.46,0.72) |
| 2006-2010 | 0.97(0.85,1.12) | 0.46(0.37,0.58) |
| 2011-2015 | 0.97(0.83,1.14) | 0.39(0.31,0.50) |
| 2016-2021 | 0.97(0.79,1.19) | 0.25(0.19,0.33) |

Note:95% CI,95% confidence internal.

**Table S6. Temporal Trends of Age-Standardized Rates for Pulmonary Arterial Hypertension in China, 1990-2050**

| Time(years) | ASIR，95%CI | | |
| --- | --- | --- | --- |
|  | Both | Female | male |
| 1990 | 0.507(0.498,0.516) | 0.519(0.508,0.531) | 0.491(0.479,0.503) |
| 1995 | 0.498(0.491,0.505) | 0.512(0.503,0.521) | 0.483(0.474,0.492) |
| 2000 | 0.490(0.483,0.496) | 0.504(0.496,0.512) | 0.476(0.468,0.484) |
| 2005 | 0.487(0.481,0.494) | 0.499(0.491,0.507) | 0.475(0.467,0.483) |
| 2010 | 0.484(0.478,0.491) | 0.491(0.483,0.498) | 0.478(0.470,0.485) |
| 2015 | 0.482(0.476,0.488) | 0.487(0.479,0.495) | 0.478(0.470,0.486) |
| 2020 | 0.491(0.484,0.497) | 0.493(0.484,0.501) | 0.486(0.478,0.495) |
| 2025 | 0.491(0.475,0.507) | 0.490(0.472,0.508) | 0.488(0.469,0.506) |
| 2030 | 0.490(0.466,0.513) | 0.486(0.460,0.512) | 0.488(0.461,0.515) |
| 2035 | 0.488(0.457,0.520) | 0.483(0.449,0.517) | 0.489(0.454,0.525) |
| 2040 | 0.487(0.448,0.527) | 0.480(0.437,0.523) | 0.490(0.446,0.535) |
| 2045 | 0.486(0.438,0.535) | 0.477(0.424,0.530) | 0.492(0.436,0.547) |
| 2050 | 0.486(0.426,0.545) | 0.475(0.409,0.540) | 0.493(0.424,0.562) |
| Time(years) | ASPR，95%CI | | |
| 1990 | 2.073(2.051,2.096) | 2.461(2.429,2.494) | 1.691(1.665,1.717) |
| 1995 | 2.107(2.088,2.126) | 2.512(2.485,2.539) | 1.713(1.692,1.734) |
| 2000 | 2.161(2.142,2.179) | 2.573(2.546,2.599） | 1.758(1.737,1.778) |
| 2005 | 2.219(2.201,2.237) | 2.620(2.594,2.646） | 1.824(1.804,1.845) |
| 2010 | 2.279(2.261,2.297) | 2.651(2.625,2.676） | 1.909(1.889,1.930) |
| 2015 | 2.318(2.300,2.335) | 2.685(2.660,2.710） | 1.950(1.929,1.970) |
| 2020 | 2.280(2.262,2.297) | 2.629(2.604,2.654） | 1.937(1.916,1.958) |
| 2025 | 2.284(2.217,2.419) | 2.622(2.539,2.705） | 1.963(1.896,2.029) |
| 2030 | 2.314(2.209,2.419) | 2.641(2.514,2.768） | 2.002(1.898,2.105) |
| 2035 | 2.344(2.203,2.484) | 2.660(2.491,2.829） | 2.041(1.901,2.180) |
| 2040 | 2.375(2.197,2.552) | 2.680(2.467,2.892） | 2.081(1.903,2.259) |
| 2045 | 2.408(2.187,2.628) | 2.701(2.439,2.964） | 2.122(1.900,2.343) |
| 2050 | 2.443(2.171,2.715） | 2.725(2.402,3.048) | 2.163(1.888,2.439) |
| Time(years) | ASDR，95%CI | | |
| 1990 | 0.612(0.593,0.632) | 0.553(0.531,0.576) | 0.721(0.686,0.755) |
| 1995 | 0.550(0.534,0.565) | 0.505(0.487,0.522) | 0.639(0.614,0.664) |
| 2000 | 0.482(0.469,0.495) | 0.451(0.435,0.466) | 0.553(0.531,0.574) |
| 2005 | 0.472(0.460,0.485) | 0.435(0.421,0.450) | 0.551(0.531,0.572) |
| 2010 | 0.603(0.589,0.616) | 0.517(0.502,0.532) | 0.742(0.720,0.765) |
| 2015 | 0.507(0.496,0.518) | 0.428(0.416,0.441) | 0.635(0.616,0.654) |
| 2020 | 0.427(0.418,0.436) | 0.379(0.368,0.390) | 0.510(0.495,0.526) |
| 2025 | 0.393(0.332,0.453) | 0.350(0.301,0.400) | 0.468(0.381,0.556) |
| 2030 | 0.359(0.271,0.447) | 0.320(0.249,0.391) | 0.430(0.302,0.557) |
| 2035 | 0.325(0.220,0.430) | 0.289(0.205,0.374) | 0.391(0.238,0.544) |
| 2040 | 0.292(0.177,0.408) | 0.260(0.167,0.354) | 0.352(0.183,0.521) |
| 2045 | 0.262(0.140,0.384) | 0.234(0.135,0.333) | 0.316(0.137,0.495) |
| 2050 | 0.236(0.110,0.362) | 0.211(0.109,0.313) | 0.283(0.099,0.467) |
| Time(years) | ASYR，95%CI | | |
| 1990 | 16.168(16.072,16.264) | 15.745(15.623,15.868) | 17.124(16.961,17.287) |
| 1995 | 14.018(13.935,14.101) | 13.883(13.774,13.992) | 14.639(14.504,14.774) |
| 2000 | 11.747(11.674,11.821) | 11.854(11.755,11.953) | 12.079(11.962,12.196) |
| 2005 | 11.019(10.950,11.087) | 11.0167(10.924,11.109) | 11.454(11.344,11.563) |
| 2010 | 13.342(13.273,13.412) | 12.641(12.548,12.733) | 14.657(14.544,14.771) |
| 2015 | 11.151(11.092,11.209) | 10.372(10.293,10.450) | 12.426(12.333,12.519) |
| 2020 | 8.925(8.878,8.973) | 8.658(8.592,8.724) | 9.562(9.488,9.635) |
| 2025 | 8.030(6.796,9.265) | 7.729(6.617,8.842) | 8.759(7.300,10.217) |
| 2030 | 7.093(5.378,8.809) | 6.790(5.261,8.320) | 7.810(5.767,9.854) |
| 2035 | 6.278(4.281,8.274) | 5.972(4.206,7.738) | 6.971(4.574,9.369) |
| 2040 | 5.566(3.399,7.733) | 5.257(3.356,7.159) | 6.227(3.605,8.848) |
| 2045 | 4.932(2.673,7.191) | 4.623(2.657,6.589) | 5.550(2.801,8.299) |
| 2050 | 4.370(2.075,6.666) | 4.067(2.084,6.050) | 4.942(2.135,7.750) |

Note: ASIR,age-standardized of incidence rate;ASPR,age-standardized ofprevalence rate;ASDR,age-standardized of death rate;ASYR，age-standardized of disability-adjusted life years;95%CI, confidence internal.
